## Supplemental data for "SARS-CoV-2 transmission dynamics in Belarus revealed by genomic and incidence data analysis"

**Supplemental Information.**

**Keywords:** COVID-19, SARS-CoV-2, Belarus, genomic epidemiology phylodynamics, effective reproduction number

°

* The authors contributed equally.

### 1 Supplemental Information

| **Model** | **Parameter** | **Prior distribution** |
|---|---|---|
| Strict clock | Clock rate | Gamma(2.56,3200) |
| HKY | Kappa<br>Gamma shape | Lognormal(1,1.25)<br>Exponential(1,0) |
| Birth-death skyline | $\mathcal{R}_e$<br>Uninfectious rate<br>Sampling proportion<br>Time of origin | Lognormal(0.8,0.5)<br>Fixed to 36.5 per year<br>Beta(1,99999)<br>Normal(0.89,0.01) for cluster 1,<br>Normal(0.919,0.01) for cluster 2 |

Table S1: BDSKY model parameters used for $\mathcal{R}_e$ estimation.

| | Before quarantine | After quarantine |
|---|---|---|
| Australia [6] | 1.63 | 0.48 |
| Russia (hospital settings) [3] | 3 | 1.76 |
| Russia (hospital settings) [3] | 3.64 | 1.85 |
| New Zealand [2] | 7 | 0.2 |
| Israel [4] | 2.1 | 0.525 |
| France [1] | 2.56 | 1.38 |
| France [5] | 3 | |
| Germany [5] | 1.75 | |
| Italy [5] | 2.4 | |
| **Belarus** (this study) | **1.95** | **1.59** |

Table S2: Estimations for the effective reproduction number $\mathcal{R}_e$ for different countries reported in the literature.

| Cluster name | Sequence Name | Country Name |
|---|---|---|
| Cluster 1 | Ukraine/203100356/2020—2020-05-28 | Ukraine |
| | Ukraine/203100318/2020—2020-06-23 | |
| | Ukraine/203100333/2020—2020-06-24 | |
| | Ukraine/203100319/2020—2020-06-23 | |
| | Ukraine/203100335/2020—2020-06-26 | |
| | Ukraine/203100336/2020—2020-06-23 | |
| | Ukraine/Rivne_55/2020—2020-06-04 | |
| | Ukraine/ChVir23535_80/2021—2021-01-10 | |
| | Ukraine/ChVir23535_23/2021—2021-01-10 | |
| | Ukraine/203100348/2020—2020-05-16 | |
| | Ukraine/Vinnytsia_12/2020—2020-05-27 | |
| | Ukraine/203100339/2020—2020-07-11 | |
| | Ukraine/ChVir23535_35/2021—2021-01-12 | |
| | Ukraine/203100320/2020—2020-06-23 | |
| Cluster 2 | Ukraine/203100361/2020—2020-04-24 | Ukraine |
| | Ukraine/Kharkiv-877/2020—2020-08-07 | |
| | Ukraine/Kharkiv-782/2020—2020-07-31 | |
| | Ukraine/Kharkiv-705/2020—2020-07-28 | |
| | Ukraine/ChVir23535_53/2021—2021-01-11 | |
| | Ukraine/ChVir23535_21/2021—2021-01-09 | |
| | Ukraine/ChVir23535_77/2021—2021-01-09 | |
| | Ukraine/ChVir23535_79/2021—2021-01-09 | |
| | Ukraine/ChVir23535_42/2021—2021-01-12 | |
| | Ukraine/ChVir23535_29/2021—2021-01-11 | |
| | Ukraine/ChVir23535_3/2021—2021-01-11 | |
| | Ukraine/ChVir23535_51/2021—2021-01-12 | |
| | Ukraine/Kharkiv-879/2020—2020-08-07 | |
| | Ukraine/ChVir23535_4/2021—2021-01-11 | |
| Cluster 1 | hCoV-19/Belarus/MN-RRCEM-Sars-Cov-2-sp-7/2021—EPI_ESL_1138983—2021-01-26 | Belarus |
| | Belarus/MI-RII-MH11337S/2020—2020-04-24 | |
| | Belarus/MI-RII-MH11351S/2020—2020-07-06 | |
| | Belarus/ChVir21882/2020—2020-10-30 | |
| | Belarus/ChVir21837/2020—2020-11-07 | |
| | Belarus/ChVir21840/2020—2020-11-30 | |
| | Belarus/ChVir21846/2020—2020-11-04 | |
| | Belarus/ChVir21895/2020—2020-10-26 | |
| | Belarus/ChVir21845/2020—2020-11-06 | |
| | Belarus/ChVir21863/2020—2020-11-04 | |
| | Belarus/ChVir21835/2020—2020-11-07 | |
| Cluster 5 | hCoV-19/Belarus/Gomel/2021—EPI_ISL_1222766—2021-02-26 | Belarus |
| | Belarus/HO-RII-MH11354S/2020—2020-11-24 | |
| | Belarus/MI-RII-MH11355S/2020—2020-11-23 | |
| | Belarus/MA-RII-MH11359S/2020—2020-10-11 | |
| | Belarus/ChVir21894/2020—2020-10-26 | |
| | Belarus/ChVir21848/2020—2020-11-09 | |
| | Belarus/ChVir21841/2020—2020-10-21 | |
| | Belarus/HO-RRCEM-MOZ11874S/2020—2020-12-08 | |

Table S3: Analyzed sequences and their sampling times.

| Cluster name | Sequence Name | Calendar Date Time of MRCA | Number of Sequences |
|---|---|---|---|
| 1 | Belarus/MN-RRCEM-Sars-Cov2-sp-7<br>Belarus/ChVir21835<br>Belarus/ChVir21863<br>Belarus/ChVir21845<br>Belarus/ChVir21895<br>Belarus/ChVir21846<br>Belarus/ChVir21882<br>Belarus/MI-RII-MH11351S<br>Belarus/ChVir21840<br>Belarus/ChVir21837<br>Belarus/MI-RII-MH11337S | April 5, 2020 | 11 |
| 2 | Belarus/ChVir21877<br>Belarus/ChVir21898<br>Belarus/ChVir21897<br>Belarus/ChVir21859 | July 9, 2020 | 4 |
| 3 | Belarus/ChVir21884 | May 4, 2020 | 1 |
| 4 | Belarus/ChVir21890 | April 4, 2020 | 1 |
| 5 | Belarus/HO-RII-MH11354S<br>Belarus/MI-RII-MH11355<br>Belarus/HO-RRCEM-MOZ11874S<br>Belarus/ChVir21848<br>Belarus/ChVir21894<br>Belarus/ChVir21841<br>Belarus/MA-RII-MH11359S<br>Belarus/Gomel/2021 | March 28, 2020 | 8 |
| 6 | Belarus/MI-RII-MH11353S | February 13, 2020 | 1 |
| 7 | Belarus/ChVir21891<br>Belarus/ChVir21892 | October 19, 2020 | 2 |
| 8 | Belarus/VI-RII-MH11358S<br>Belarus/HM-RRCEM-Sars-CoV2-sp-1 | September 27, 2020 | 2 |
| 9 | Azerbaijan/RRCEM-sp_3/2021 | April 30, 2020 | 1 |
| 10 | Belarus/ChVir21888 | April 30, 2020 | 1 |
| 11 | Belarus/ChVir21832 | April 30, 2020 | 1 |
| 12 | Belarus/ChVir21843 | April 30, 2020 | 1 |
| 13 | Belarus/ChVir21842 | April 8, 2020 | 1 |
| 14 | Belarus/ChVir21889 | April 8, 2020 | 1 |
| 15 | Belarus/MI-RRCEM-Sars_Cov_Vis_68/2021 | April 5, 2020 | 1 |
| 16 | Belarus/ChVir2073 | January 30, 2020 | 1 |
| 17 | Belarus/ChVir21878 | April 8, 2020 | 1 |
| 18 | Belarus/ChVir2072<br>Belarus/ChVir2070 | February 21, 2020 | 2 |

Table S4: Inferred clusters and their times of MRCA.

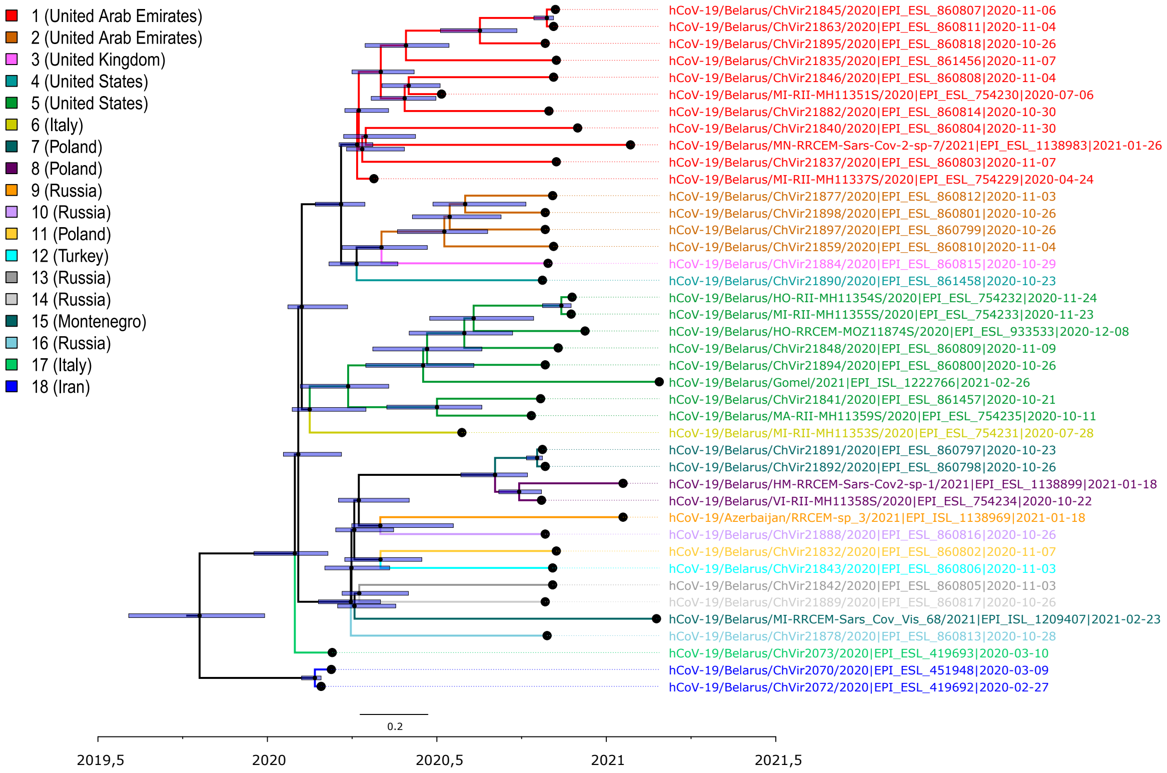


Figure S1: The annotated maximum clade credibility tree with sequence names and visualized 95 HPD in blue: clusters/local lineages numbered from one to eighteen, tree branches and sequence names color-coded by cluster IDs; cluster sources added in parentheses.

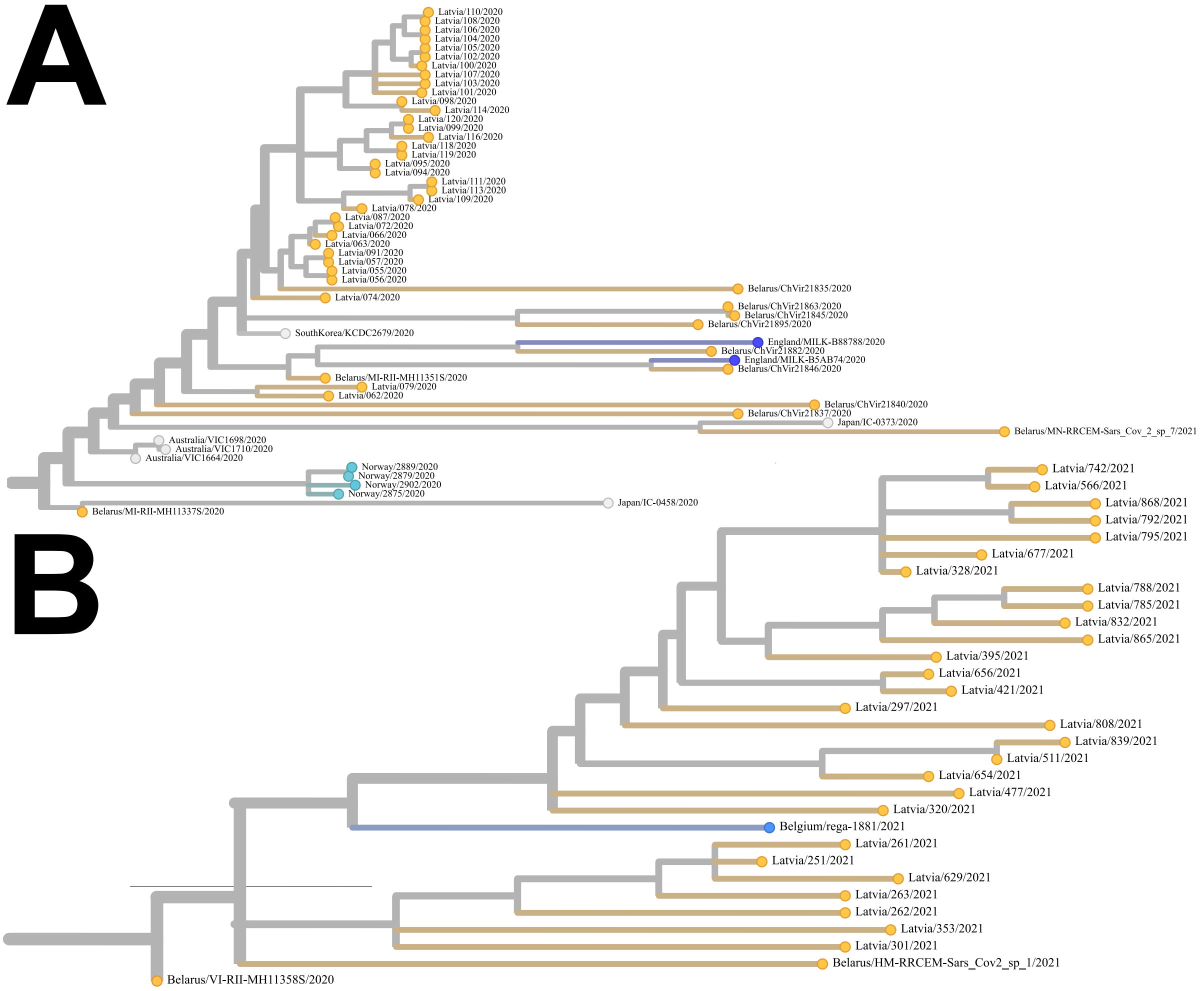


Figure S2: Latvian lineages originated from two alleged introductions from Belarus. Trees were visualized by Nextstrain
